# The risk of Asthma hospitalisation associated with proximity to the liquefied natural gas Ports in Gladstone, Australia

**DOI:** 10.1101/2025.10.01.25337041

**Authors:** Thuy Lam

**Affiliations:** School of Public Health and Social Work. Queensland University of Technology

## Abstract

The mining of natural gases has been ongoing for several decades, along with on-going research effort on the impact it may have on human health. Despite the research effort invested into this issue, a knowledge gap still exist on how extensive extractive industry emissions can impact the health of populations living in proximity to these mining activities. This study aims to investigate the risk of asthma hospitalisation associated with proximity to the liquefied natural gas (LNG) ports in Gladstone, Australia. A total of three LNG terminals were identified on Curtis Island that were set as the exposure. The study area was established using a 100km circular buffer around the exposure, which encapsulated a total of 167 suburbs. The asthma hospitalisation rate was calculated for each of the 167 suburbs and sociodemographic data was collated using the Socio-Economic Indexes for Areas (SEIFA), this includes the Index of Relative Socio-Economic Advantages and Disadvantages (IRSAD), the Index of Education and Occupation (IEO), and the Index of Economic Resources (IER). Data collected for the suburbs in Gladstone were all tested for spatial autocorrelation. Spatial autocorrelation analysis was performed using GeoDa, implementing a Global Bivariate Moran’s I and Local Moran analysis. The results of this study have identified that a lack of diverse variable incorporation or multivariable analysis can greatly impact the implementation of spatial and temporal analyses. The study conclude that the use of spatial statistics is highly recommended for future studies as incorporating spatial analysis can create a more comprehensive visualisation of high-risk areas. Additionally, the incorporation of a multivariable spatial analysis can potentially benefit prevalence modelling in complex scenarios concerning multiple variables that can impact determinant of disease in the general population.

## Background

Liquefied natural gas (LNG) is a commodity derived from the rapid cooling of natural gas. As of 2022, Australia was ranked second for LNG export and seventh in global LNG production. In 2022 LNG exports were of record levels, recording an estimated 81.4 million tonnes of gas exported to international markets, this equates to 70% of Australia’s natural gas production according to Geoscience Australia (2024). In the global energy market, LNG is regarded as one of the fastest increasing fossil fuel due to its low emission, high energy production potential, price and ease of transportation over long distance and durations (Reserve Bank of Australia, 2011). Despite the high demands for LNG in the global market, the production of LNG involves an energy intensive industrial process. To produce LNG, natural gas is extracted through boreholes that is created by drilling wells, typically 200 – 1000 metres from the land surface into the coal seam to extract coal seam gas (CSG). Wells drilled into the coal seams extracts CSG and other byproduct like wastewater containing naturally occurring radioactive elements, arsenic, mercury and high salinity waste water that will require further treatment for that waste water to become reusable or safely disposed as waste (Haswell et al., 2023). In Queensland, extracted CSG is transported through an extensive network of pipeline to the LNG plant, where the gas is processed (Bahadori, 2014).

The production of LNG typically involves four stages, pretreatment, acid gas removal and dehydration, removal of heavy hydrocarbons and finally the separation and liquefaction. At the pretreatment stage, extracted CSG will undergo dust, slug (water and condensation), sulphide (H_2_S) and mercury (Hg) removal as these byproducts can cause corrosions and freezing issues once CSG is passed through the aluminium heat exchangers. At the second stage, carbon dioxide (CO_2_) is absorbed and removed from CSG using an amine absorber, whilst water is removed by another absorber. This process ensures that further impurities is removed to prevent ice formation issues in the subsequent liquefaction process (Mokhatab & Economides, 2006). The third stage involves the removal of heavy hydrocarbons (C5+) through fractionation and precooling to roughly -35℃ by propane. The separation and liquefaction is the final stage in the production of LNG, whereby purified coal seam gas will be passed through channels of mixed refrigerant (MR) to be cooled down. At this stage cooling and separation of vapor and liquid happens simultaneously. Through different sections of the cooling process, the streams is controlled to travel at pressure to further reduce the temperature of the gas to induce liquefaction of the natural gas. At the final cooling process, the natural gas is typically liquefied and sub-cooled to temperatures between -150 ℃ to -162℃. Once processed LNG is transferred into storage tanks awaiting transportations (Bahadori, 2014). It is during these LNG production processes that occur at the LNG terminal, that creates the most air pollution as flaring is a part of LNG production.

During the production of LNG, the flaring operations and emissions from these LNG plants can be a great cause of concern to human health impacts (Anejionu et al., 2015). Emissions from the LNG plant specifically, can include nitrogen oxide (NO_x_), carbon monoxide (CO), carbon dioxide (CO_2_), methane, nitrous oxide (N_2_O), sulphur dioxide (SO_2_), particulate matters (PM), and volatile organic compounds (VOCs). These chemical components found in LNG plant emission, are known to impact human health through various routes, with some chemical components like benzene is a known carcinogen. Long-term exposures to these substances have been reported to impact health, increase hospitalizations and reduce life expectancies specially regarding respiratory health, heart disease, cancer and the reproductive system (Environmental Health Project, 2023). Emissions from LNG plants typically occur during daily operations like flaring and venting, yet despite these negative implications to public health, emissions from LNG plants is difficult to quantify due to their intermittent activities and lack of adequate monitoring and reporting (Tran et al., 2024). Reporting of flaring operations in recent time has also been placed under high scrutiny due to the underestimation of flaring requirements in gas plants. According to Tran et al (2024), there is a gap in the knowledge regarding the characteristics of flaring-related emissions and their contributions to local air quality in a real-world setting (2024). Furthermore, this knowledge gap is further influenced by factors such as the waste gas composition, operating conditions, combustion completeness/efficiency and local meteorological condition (Tran et al., 2020). This process have remained difficult to track and record as a review into the Gladstone LNG terminals have shown that in recent years several infringement notices have been issued to the LNG terminals in Gladstone over issues relating to record-keeping obligations, failure to report an oil spill in the Great Barrier Reef World Heritage Area and water turbidity issues near Curtis Island World Heritage reserve site (Santos, 2023).

Extensive research have shown that air pollution is highly associated with many high-risk conditions relating to human respiratory health, like asthma, heart disease, and cancer reproductive-related disease (Cushing et al., 2021; Environmental Health Project, 2023). Although asthma can be associated to numerous natural sources of environmental impact, it was found that asthma is also correlated with industrial-related processes and is mainly related to pollutants categorised as gaseous and particulate matter (Khreis et al., 2019; Manisalidis et al., 2020; Veremchuk et al., 2018). As a chronic inflammatory condition, asthma is characterized with symptoms such as dyspnoea, wheezing, coughs, and chest tightness due to restricted air flow (Tiotiu et al., 2020). Furthermore, evidence have suggested that air pollution can not only aggravate existing asthma symptoms, but it can also lead to new onset of asthma (Buonocore et al., 2023; Huff et al., 2019). Exceeding levels or prolonged exposure to O_3_ generated from the chemical reactions between the gases emitted through flaring operations and VOCs have the potential to induce oxidative stress resulting in airway inflammation, and reduced lung functions in asthmatic individuals (Anenberg et al., 2018; Niu et al., 2018). Despite this research, there remains a knowledge gap on how extensive extractive industry emissions can impact the health populations in populations living in proximity to these activities (Cortes-Ramirez et al., 2019).

These knowledge gaps is further challenged as multiple variables or confounding factors can influence how impact is interpreted or presented in a real-world. Variables like, proximity to LNG port, flaring tendency, combustion efficiency, pollution levels, population concentrations and exposure durations can all have the potential to determine how human health is impacted. Therefore, the aim of this research is to determine if there is an association between asthma hospitalization and the patient’s proximity to the LNG plant. The studied area is located on Curtis Island, Gladstone, Queensland, home to the three largest LNG terminals in Queensland, which also is the only LNG terminals that are in close proximity to the residents of Gladstone.

## Methods

### Study area and data source

The study area includes suburbs within Gladstone, Queensland and Curtis Island which is where the three major LNG plant/terminal is located (figure 1). Gladstone is located at latitude -23.8490, longitude 151.2630 and the population in Gladstone as of the 2016 census data is 61,640 people. There is a total of three LNG terminals in Gladstone, all of which is located on Curtis Island, therefore, to ease the complexity of the data visualisation process on the geographical analysis tool the LNG terminal located at the centre of the three LNG terminal was chosen as the centre-point to establish the circular buffer (Queensland Curtis LNG Terminal), the LNG plant is located at latitude - 23.7693, longitude 151.199 portside on Curtis Island. A 100km circular buffer was considered as the starting buffer size, since studies of association between port proximity and health impact is limited, and a 100km buffer will also cover more suburbs creating a more substantial dataset. The established 100km buffer covered a total of 167 suburbs in Gladstone and suburbs that is not completely within the 100km buffer boundary is included in the count, if the centroid (centre-point) of the suburb is within the 100km buffer boundary.

**Figure 1.**
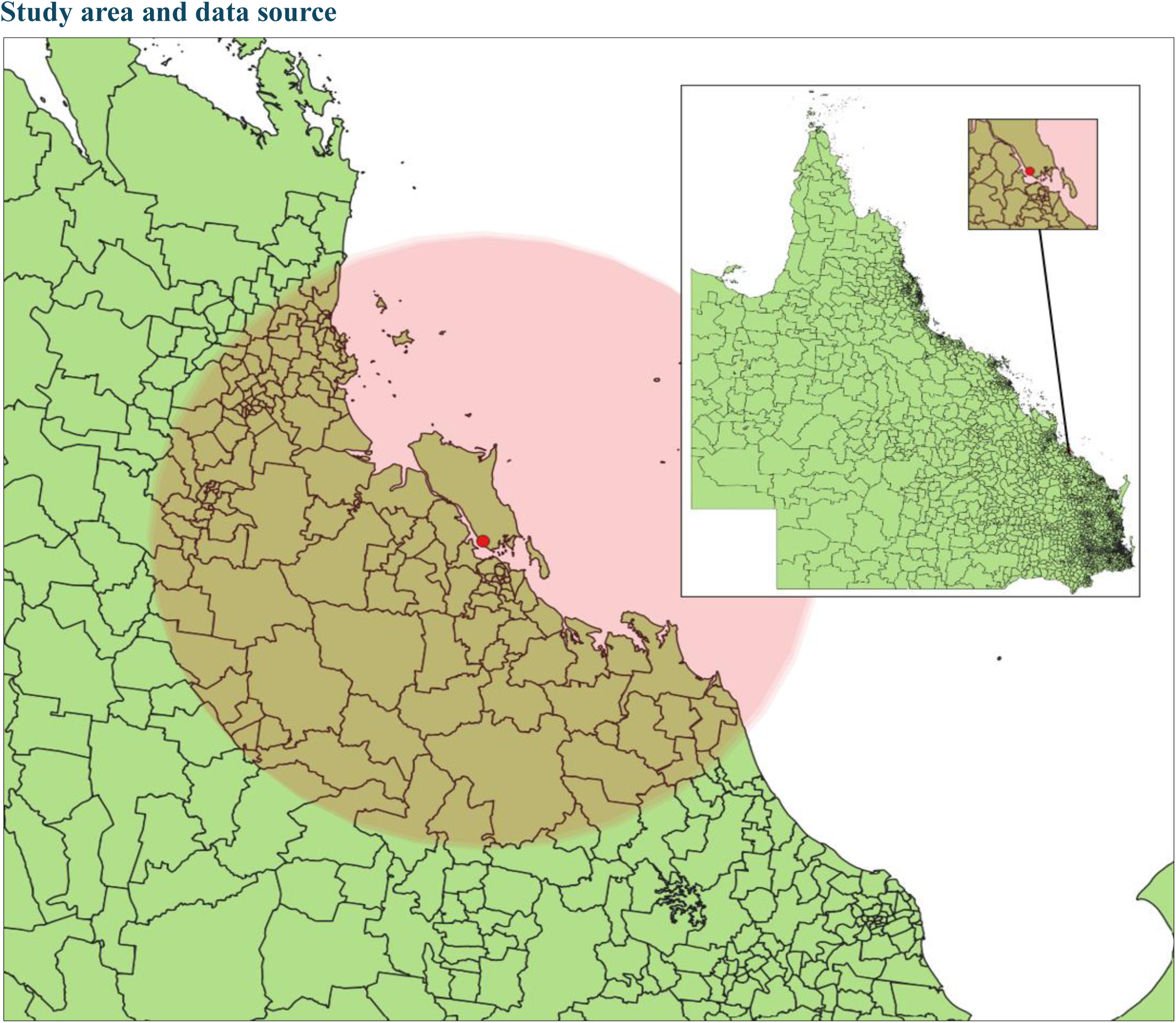
Study area: Gladstone, Queensland Australia and the location of the liquefied natural gas (LNG) port (red marker) as well as the coverage of the 100km buffer.

Demographic data, for Gladstone was collected using the Table Builder function from the Australian Bureau of Statistics (ABS). To generate data relative to population size and socioeconomic circumstances of the population within the studied area, the 2016 census data was retrieved from the Australian Beau of Statistics (ABS) using the provided Table Builder function. To build the dataset of interest the Table Builder function was utilised along with several filters. Using the pro setting, the 2016 census data was first filtered for the type of population count. The type of population count utilised was the place of usual residence, and this was further filtered to geographical area, which was set as state suburbs (SSC). The final filter was set as Queensland as it is the study area. The dataset was built to contain several variables of interest, and this include, population size, sex, and the three socioeconomic indexes. The Socio-Economic Indexes for Areas (SEIFA) include four socioeconomic indicators developed by the ABS to measure relative socio-economic advantages and disadvantages based on decile for each geographical area in Australia. The SEIFA data is based on the socioeconomic information obtained from the five-yearly census. Only three of these indexes will be utilised. This includes the Index of Relative Socio-Economic Advantages and Disadvantages (IRSAD), The index of Education and Occupation (IEO) and lastly, the Index of Economic Resources (IER). The addition of the SEIFA data aid in determining if there is a positive correlation between socioeconomic advantages and disadvantages to health and educational outcomes. To obtain the SEIFA data, the statistical area was set at level 2 (SA2).

### Spatial analysis software and Statistical software

To generate spatial information and visualisation of spatial data relating to the study area, a geographical information system was utilised. QGIS (version 3.38.1) was utilised as part of the mapping and layering tool for this project. Through the use of QGIS, a shapefile for Queensland, Australia was set as the base map, and any additional data was included in the form of a layer on top of the Queensland basemap. The LNG port locations and surrounding suburbs was identified to aid in visualising the buffer points and suburbs within the 100km buffer. As a feature included in the QGIS software, each layer added onto the base map of Queensland, was set as either centroids, buffer or polygon and is further differentiated by colour code for ease of visualisation. Demographic data including population size, sex, SEIFA and asthma hospitalisation rate for each suburb in Queensland was load as one dataset on top of the Queensland basemap and was filtered according to each stage of the projects requirement. Further analysis of the dataset was carried using a software called GeoDa (version: 1.22.0.4), and this software was utilised as a mean to conduct spatial data analysis and spatial autocorrelation analysis.

The spatial autocorrelation analysis was carried in GeoDa using two types of analysis method, which includes the Global Bivariate Moran’s I and Local Moran. The purpose of running a Global Bivariate Moran’s I is to analyse the spatial autocorrelation for the entire dataset which will generate a single Moran’s Index value. Bivariate Local Moran was implemented to evaluate spatial autocorrelation of similar locations to their neighbours and to generate a risk estimate for each location. These two test were both utilised as the Global Moran’s I can indicate if there is clustering evidence in the dataset, whilst the Local Moran can visually indicate the location of the clusters and their significances. The Global Bivariate Moran I is calculated in GeoDa using the following equation:

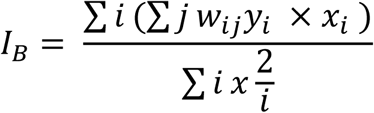

where, *I*_*B*_ is the bivariate Moran’s *I* statistics, *x*_*i*_ is the value of the variable at location *i*, and *w*_*ij*_ is the spatial weight that will determine the relationship between *i* and *j*, whilst *y*_*i*_ represents the observed value of the variable of interest at location *i*. To carry out any spatial autocorrelation analysis, a spatial weight was created, and this spatial weight created in GeoDa contains information regarding the structure of each neighbouring suburb. There are several types of contiguity weight to choose, however the Queen contiguity weight was utilised for the spatial autocorrelation analysis as the Queen contiguity weights can yield more neighbours based on common vertices rather than edges, which is ideal for geographical data that have irregular polygons. The second spatial autocorrelation analysis carried out was the Bivariate Local Moran’s I. Using GeoDa the results obtained were generated based on the following equation.

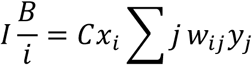

where *Cx*_*i*_ represents the spatial lag of *x*_*i*_, which is the value of the variable at location *i*.

These two analyses were carried out using the same Queen contiguity weight and a Moran scatter plot was generated as part of the Global Bivariate Moran’s I analysis. Additionally, a cluster map and significant map were created for the Bivariate Local Moran’s I analysis. Dataset visualisation and multivariable correlation analysis were carried out using R studio (version: 1.2.5033) (R Core Team, 2024). The same dataset was utilised across the three software in this project. Using R studio and the packages tidyverse (Wickham H et al., 2019), ggplot2 (Wickham H, 2016), and sjPlot (Lüdecke D, 2024), several analyses were carried out. This include a multivariate correlation analysis using the Pearson’s method and graph visualisation was carried out using ggplot2.

### Asthma Hospitalisation data

Hospitalisation data was obtained from the Queensland Hospital Admitted Patient Data Collection. The hospitalisations primary diagnosis was categorised according to the main chapters of the International Classification of Diseases version 10, Australian Modification (ICD-10-AM) to extract hospitalisation records with ICD code J.45 (asthma). The analysis of these data was approved by the Children’s Health Queensland Hospital and Health Service Human Research Ethics Committee (HREC/21/QCHQ/80236, 21/10/2022)

Using the asthma hospitalisation data obtained from Queensland’s Health, the asthma hospitalisation rate per 1000 people-year was calculated. The following equation was used to calculate the hospitalisation rate:

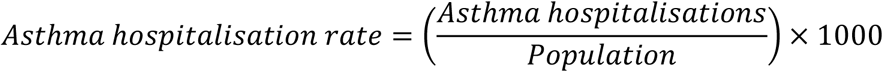

## Results

There were a total number of 3264 suburbs identified in Queensland (Table.1), of which only 167 suburbs were within the 100km buffer that was established for this study.

Figure 2 shows the asthma hospitalisation rate per 1000 people annually for the female population is much greater than the male population, with a noticeable spike in hospitalisation rate in 2017. In 2020 the hospitalisation rate for the male and female population seems to be similar to each other.

**Figure 2.**
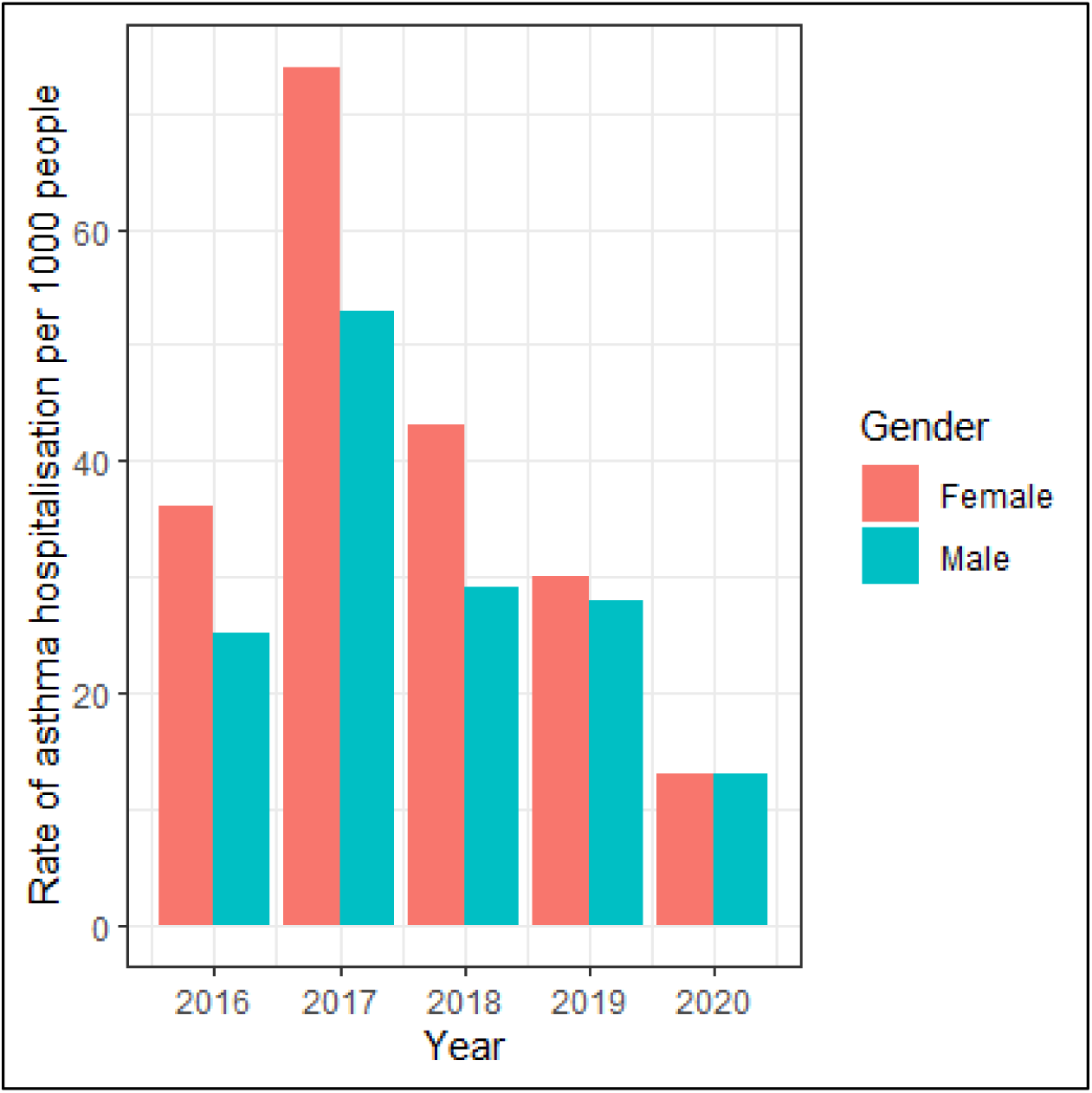
Asthma hospitalisation rate per 1000 people-year across 5 years for the male and female population.

Additionally, there are no escalating trends noted across each year.

The results obtained from the multivariable correlation analysis for female over the period of 5 years (Table 2) show that there is no positive correlation between the female population and any of the five variables tested (Distance, Male, IRSAD, IER, IEO). This negative correlation is seen across the five years with very minimal changes.

**Table 1.** Characteristics of the data included in the analysis.

|  | Total |
| --- | --- |
| Total number of suburbs in QLD | 3264 |
| Total number of suburbs within 100km buffer | 167 |
| Total number of suburbs that has asthma hospitalisation records | 86 |
| Total number of suburbs with zero population | 162 |

**Table 2.** Multivariate correlation analysis for Female.

|  | 2016 | 2017 | 2018 | 2019 | 2020 |
| --- | --- | --- | --- | --- | --- |
| **Distance** | 0.443 (<.001) | -0.067 (.542) | -0.072 (.510) | -0.021 (.851) | -0.150 (.171) |
| **Male** | 0.841 (<.001) | 0.913 (<.001) | 0.989 (<.001) | 0.845 (<.001) | 0.628 (<.001) |
| **IRSAD** | -0.199 (.067) | -0.186 (.089) | -0.190 (.082) | -0.145 (.185) | -0.099 (.368) |
| **IER** | -0.287 (.008) | -0.286 (.008) | -0.287 (.008) | -0.289 (.007) | -0.166 (.128) |
| **IEO** | -0.063 (.565) | -0.054 (.624) | -0.052 (.635) | 0.016 (.888) | -0.098 (.372) |

*Note. Computed correlation using Pearson-method with listwise-deletion for the female population within the 100km buffer of the liquefied natural gas (LNG) Port in Gladstone, Queensland. IRSAD: Index of Relative Socio-Economic Advantages and Disadvantages, IER: Index of Economic Resources, IEO: Index of Education and Occupation.*

**Table 3.** Multivariate correlation analysis for Males.

|  | 2016 | 2017 | 2018 | 2019 | 2020 |
| --- | --- | --- | --- | --- | --- |
| **Distance** | 0.006 (-0.953) | -0.057 (.603) | -0.216 (.047) | -0.152 (.166) | -0.163 (.135) |
| **Female** | 0.841 (<.001) | 0.913 (<.001) | 0.898 (<.001) | 0.845 (<.001) | 0.628 (<.001) |
| **IRSAD** | -0.191 (-0.08) | -0.138 (.209) | -0.090 (.411) | -0.166(.128) | -0.249 (.022) |
| **IER** | -0.279 (-0.01) | -0.224 (.040) | -0.212 (.051) | -0.293 (.006) | -0.325 (.002) |
| **IEO** | -0.014 (.901) | -0.028 (.797) | -0.033 (.763) | -0.061 (.578) | -0.165 (.132) |

*Note. Computed correlation using Pearson-method with listwise-deletion for the male population within the 100km buffer of the liquefied natural gas (LNG)Port in Gladstone, Queensland. IRSAD: Index of Relative Socio-Economic Advantages and Disadvantages, IER: Index of Economic Resources, IEO: Index of Education and Occupation.*

To identify if the proximity to an LNG terminal is correlated to asthma hospitalisation, five scatterplots with trendlines were generated (Figure 3) over 5 years. The data was further segregated according to gender. As shown in figure 3, the x-axis is graphed using the proximity to the LNG terminals and the y-axis is graphed using the asthma hospitalisation rate per 1000 people annually. A visual inspection of the graph shows that there is a trend amongst the 5 years. The trend shows that the further away or the greater the distance is between the suburbs and the LNG terminal, the smaller the incidence of asthma hospitalisation is recorded. The trend line also shows the same tendency, despite the presence of several outliers. The outliers is present in each of the 5 years, however there is a tendency for outliers to be in greater concentrations at distance closest to the LNG terminals and furthest away from LNG terminals. Very limited number of outliers is seen at between the 37km and 67km locations. The datapoint is also most concentrated at two locations, and this seen at the distance closest to the LNG terminals and the distance furthest away from the LNG terminals. The graphs in Figure 3 also shows that female have a higher rate of asthma hospitalisation than male across the 5-year survey, with the greatest difference seen in the year 2016.

**Figure 3.**
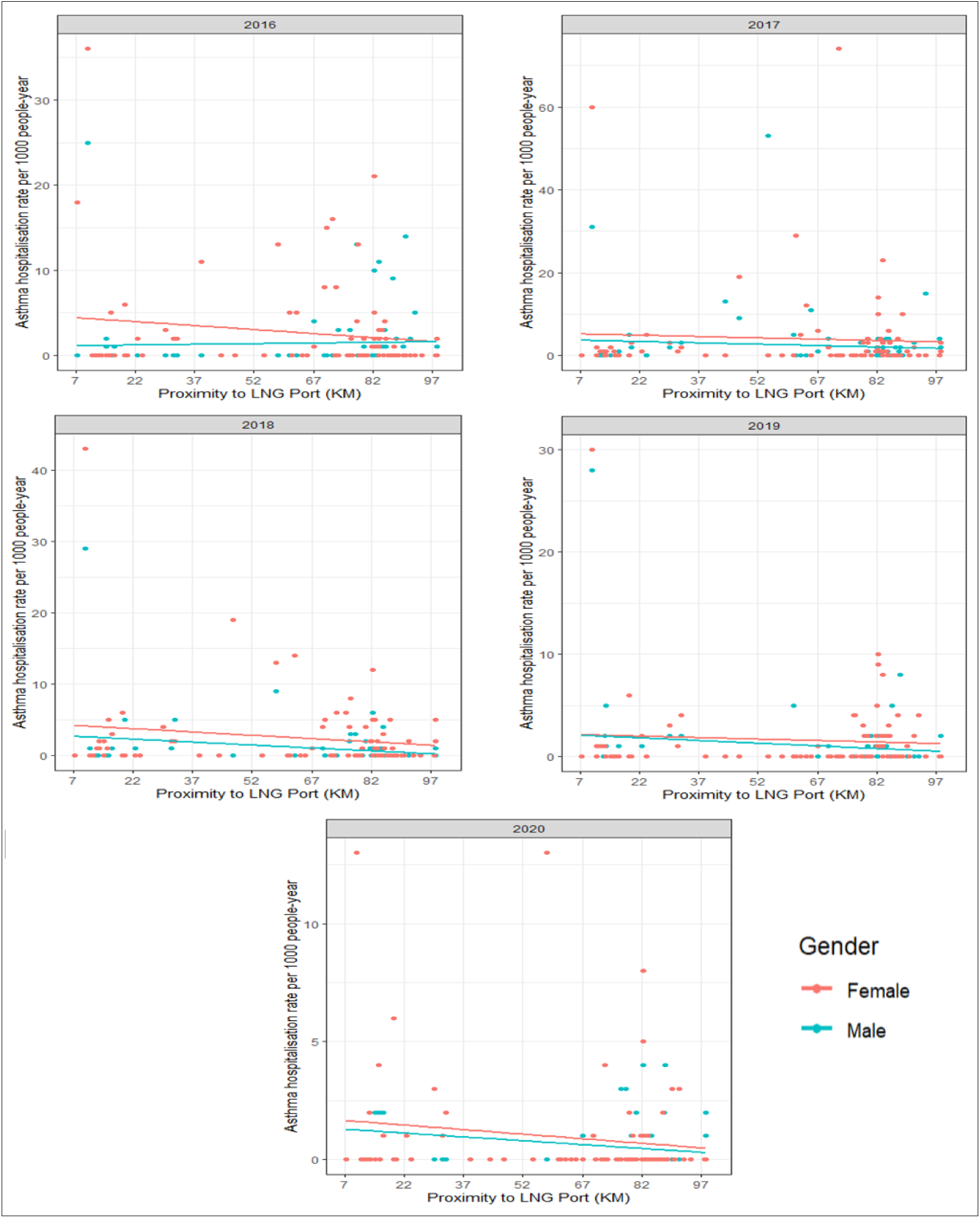
Asthma hospitalisation rate per 1000 people annually (across 5 years) according to their proximity to the LNG terminals.

The results obtained from the multivariable correlation analysis for female over the period of 5 years (Table 2) show that there is a positive correlation between distance and female asthma hospitalisation. Additionally, no positive correlation was found between the female population and any of the other four variables tested (Male, IRSAD, IER, IEO). This negative correlation is seen across the five years with very minimal changes.

A multivariable correlation analysis was also carried out for the male population, based on the same variables measured in the female population. The results (Table.3) show that there is no positive correlation between the male population and the variables, and this negative correlation remained unchanged across the 5-year period.

The results in Figure 4, show the decile ranking of each suburb according the IRSAD, relative to their proximity to the LNG terminal and the rate of asthma hospitalisation rate per 1000 people annually. A trend can be identified across the 5-year survey, with decile 5 and decile 2 appearing in higher frequency the closer it is to the proximity of the LNG terminal. The decile ranking is from decile 1 to decile 10, the smaller the decile the more disadvantage and the higher the decile the less disadvantage they are. Figure 4 show that the least disadvantage of the group, those in decile 7 is furthest away from the LNG terminals, whilst those in decile 4, the middle decile is typically located at the mid-way section. The most disadvantaged, those on decile 1 is also located at the furthest distance from the LNG port. Those in decile 5 is also appearing in greater numbers when compared to another decile.

**Figure 4.**
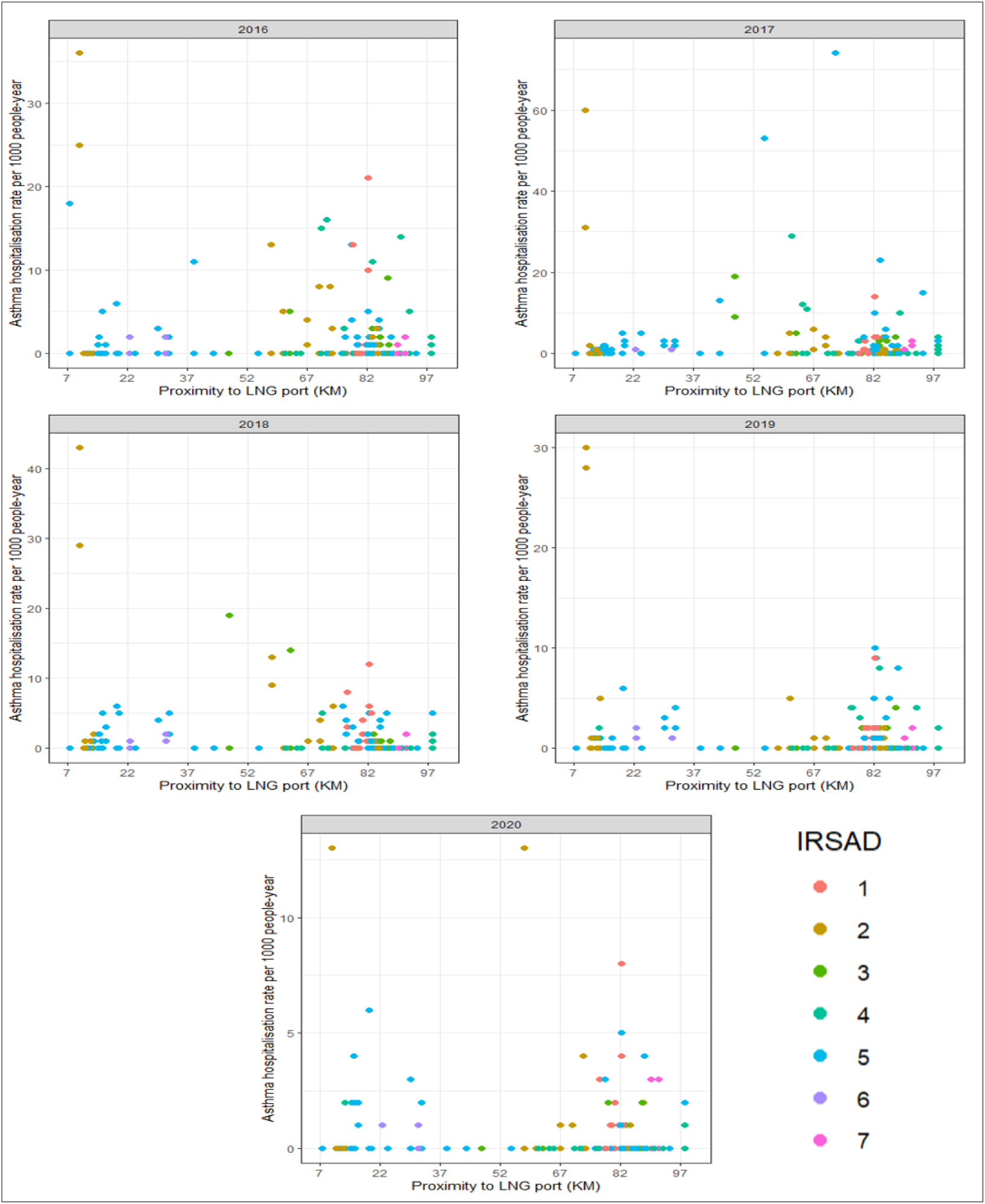
Index of Relative Socioeconomic Advantages and Disadvantages (IRSAD) in suburbs within the 100km buffer of the LNG terminal.

Figure 5, show the results for the Index of Economic Resources (IER). Across the 5-years. the lack of economic resources or those in decile 1 is typically found in proximity closest to the LNG terminal. Although the numbers is small, but those in decile 10 being the least lacking economic resources is found in distance furthest away from the LNG terminals. Those in decile 8 is mainly concentrated around the 7km and 50km mark of the 100km buffer. Decile 8 is not seen in distances further away from the LNG terminals. The outliers seen in Figure 5 is also commonly those from decile 1, with only a slight difference seen in 2017 and 2020, where there were outliers from other deciles.

**Figure 5.**
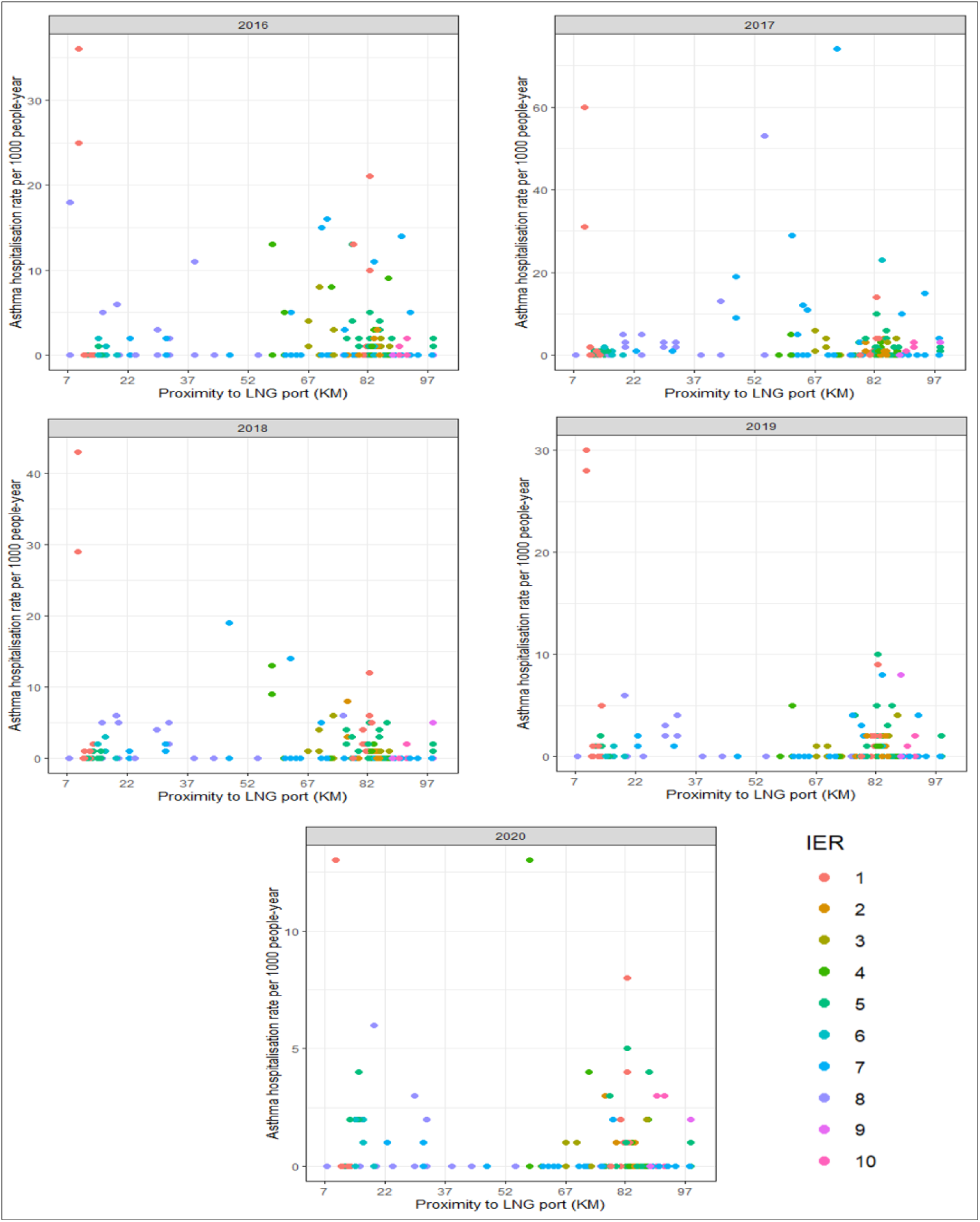
Index of Economic Resource (IER) in suburbs within the 100km buffer of the LNG terminal.

Figure 6 show that those closer to the LNG terminals tends to be in deciles 2 and 3. This tendency is seen across the 5 years. Those in decile 1 is only seen at the midway point of the buffer and scattered out towards the 100km location. The furthest outliers in each year tend to be those from decile 2. Only in 2017 and 2020 there were other deciles in the outliers.

**Figure 6.**
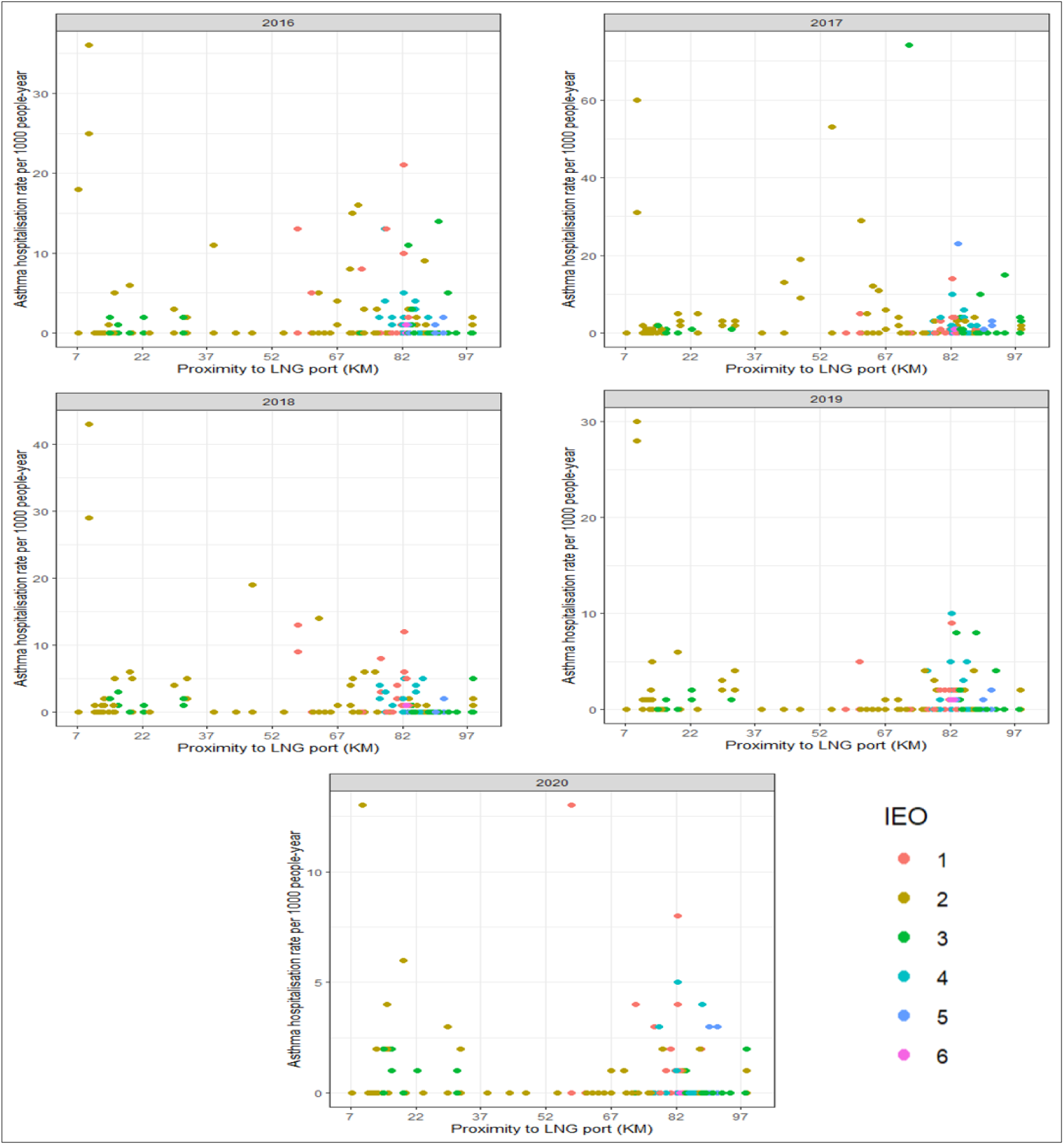
Index of Education and Occupation (IEO) in suburbs within the 100km buffer of the LNG terminal.

Figure 7 show the distribution of asthma hospitalisation rate per 1000 people annually relative to the location of the LNG terminal (red centroid). The colour gradient on the map is an indicator for rate of asthma hospitalisation in each suburb, and the gradient indicates that female asthma hospitalisation rate (right) is different to the male asthma hospitalisation rate (left) as darker gradient is more prevalent in the female group. However, the distribution pattern is relatively similar, with most cases distributed in close proximity to the LNG terminals. The largest rate of asthma hospitalisation is more prevalent in suburbs with larger population counts while smaller population record smaller rates of asthma hospitalisation.

**Figure 7.**
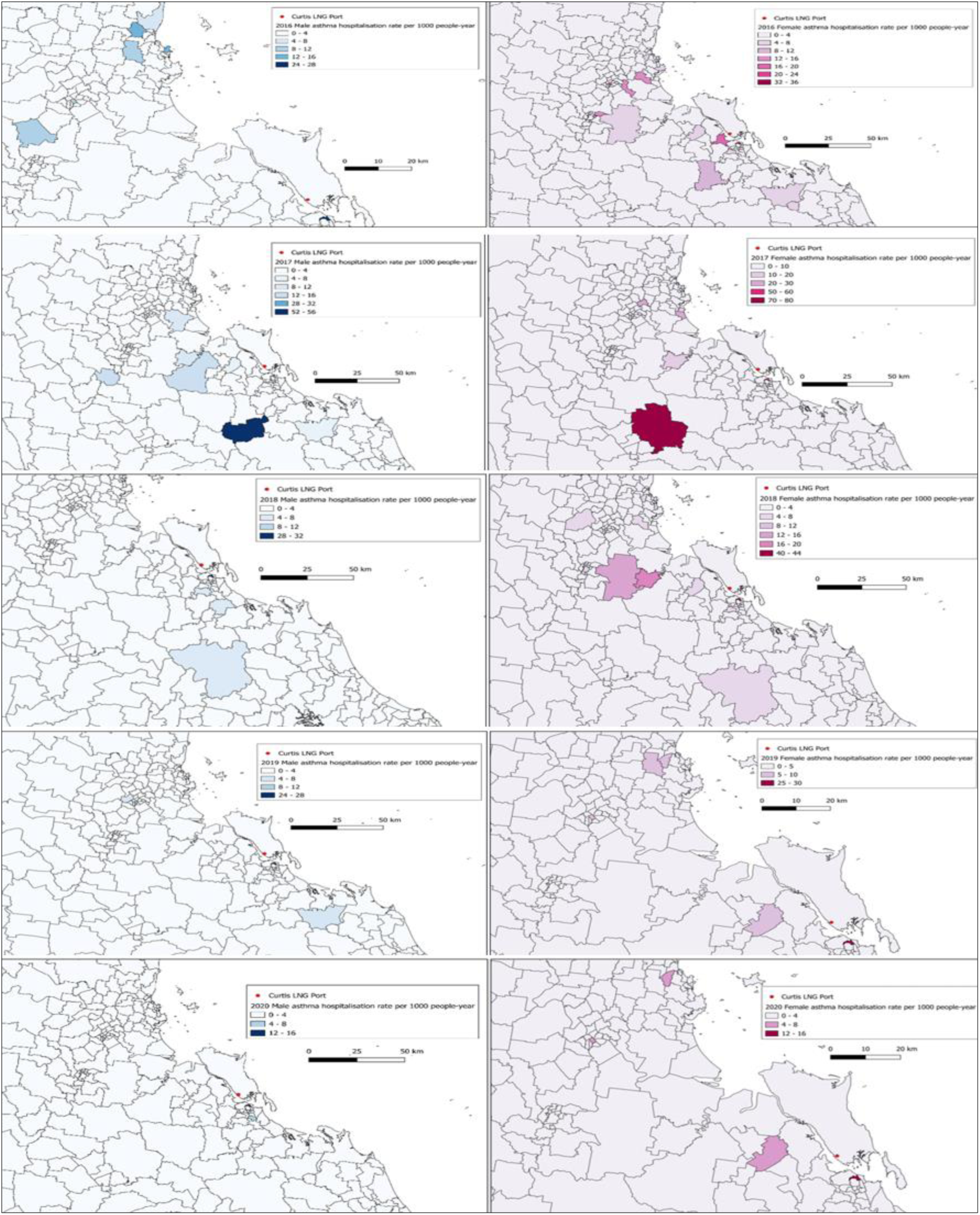
Distribution of asthma hospitalisation rate per 1000 people annually, separated by male (left) and female (right), over a period of 5 years.

Figure 8 shows the Global Bivariate Moran I analysis, using the two variables the asthma hospitalisation rate per 1000 people annually and the proximity to LNG terminals. The analysis was carried out each year, using the combine asthma hospitalisation rate for both sexes. The result from the analysis shows that there were varying degrees of positive spatial autocorrelations in each year as there is evidence of clustering.

**Figure 8.**
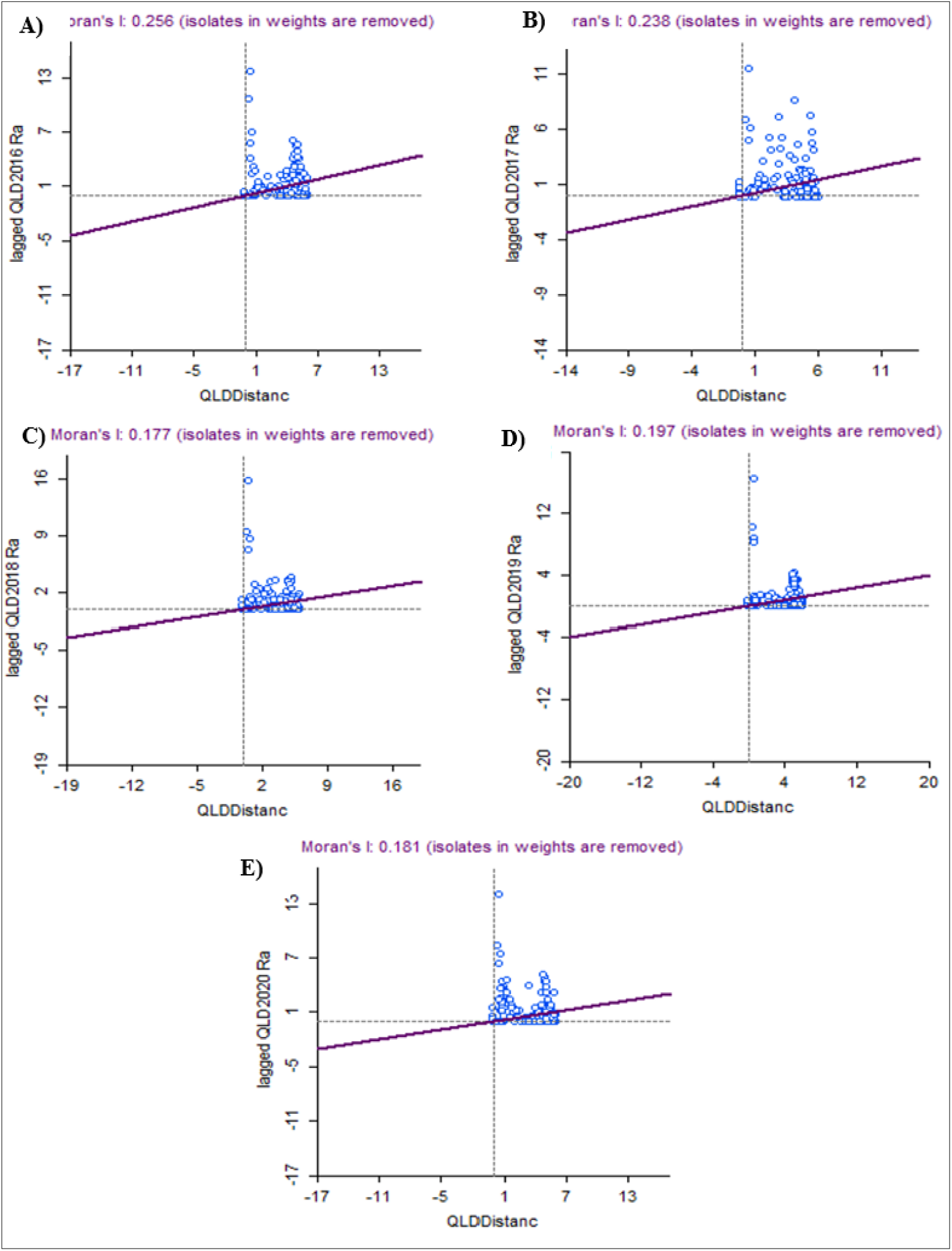
Bivariate Global Moran’s I analysis over a period of 5 years using the annual rate of asthma hospitalisation per 1000 people and the proximity to LNG terminals.

The evidence of positive spatial association can be observed across all analysis for the 5-year period, by observing the position of datapoint on the Moran’s scatterplot. The Moran’s scatterplot is divided into four quadrants, from left to right the quadrants can be referred to as low-high (negative association), high-high (positive spatial association), low-low (positive spatial association) and high-low (negative association). As the results in figure 8 shows that the datapoints all remained within the high-high quadrant, it shows that there is a significant level of positive spatial autocorrelation, therefore there is confidence that similar asthma hospitalisation rate is observed in the surrounding locations. The year with the highest spatial autocorrelation record is 2016 (I = 0.256) and 2017 (I = 0.238). The smallest Moran’s Index out of the 5 years is the year 2016 (I = 0.177), with outliers evidently separated from the larger clusters. All 5 years has outliers to varying extent; however, the outliers do not extend into other quadrants. This analysis is different to the multiple variable correlations performed in R studio, as this incorporates spatial distribution factors like proximity between neighbouring areas of similar and dissimilar values, which will generate different statistical outcomes.

A Bivariate Local Moran I’s analysis was carried over a period of 5 years from 2016 to 2020, to assess the locations of the clustering. Using the combined rate of asthma hospitalisation of both sexes, Figure 9 shows that there is evidence of high-high clustering. In 2016, 102 suburbs recorded high-high clustering, and this increased in 2017 with 118 suburbs with high-high clustering. The drop in suburb numbers with high-high clustering, starts to appear from the year 2018 to the year 2020. Significant map (Figure 9) also supports the clustering map, suggesting a high confidence of spatial autocorrelation amongst the suburbs that is in closer proximity to the LNG terminals.

**Figure 9.**
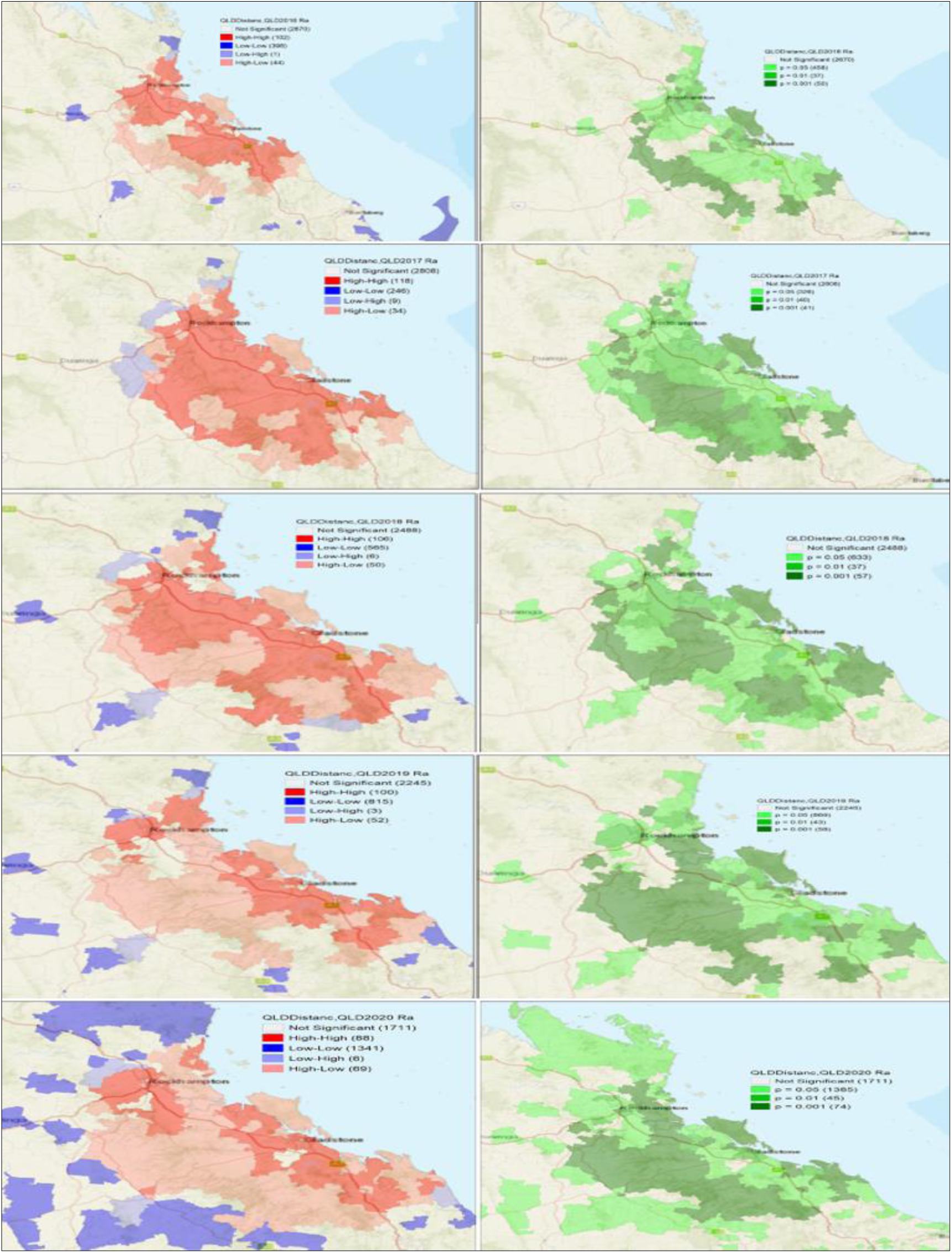
Bivariate Local Moran’s I analysis – Cluster map (left) and significant map (right) over a period of 5 years from 2016 to 2020.

Figure 10 show the Bivariate Local Moran’s I analysis for the three SEIFA tested against the proximity to the LNG port. Figure 10 show that each SEIFA displays different clustering patterns. The IRSAD only recorded 9 suburbs with high-high clusters very few is located where the LNG terminals is located. The IER analysis obtained more high-high clusters with a total of 49 suburbs, despite many low-low suburbs, the high-high clusters were predominantly within closer proximity to the LNG terminals. The IEO recorded zero high-high suburbs, and only high-low clusters with a total of 65 suburbs, with greater numbers seen in the low-high and low-low measures. Despite the differences in the clustering patterns of each SEIFA, there is still evidence to suggest a high degree of spatial autocorrelation between SEIFA and the proximity to an LNG terminal.

**Figure 10.**
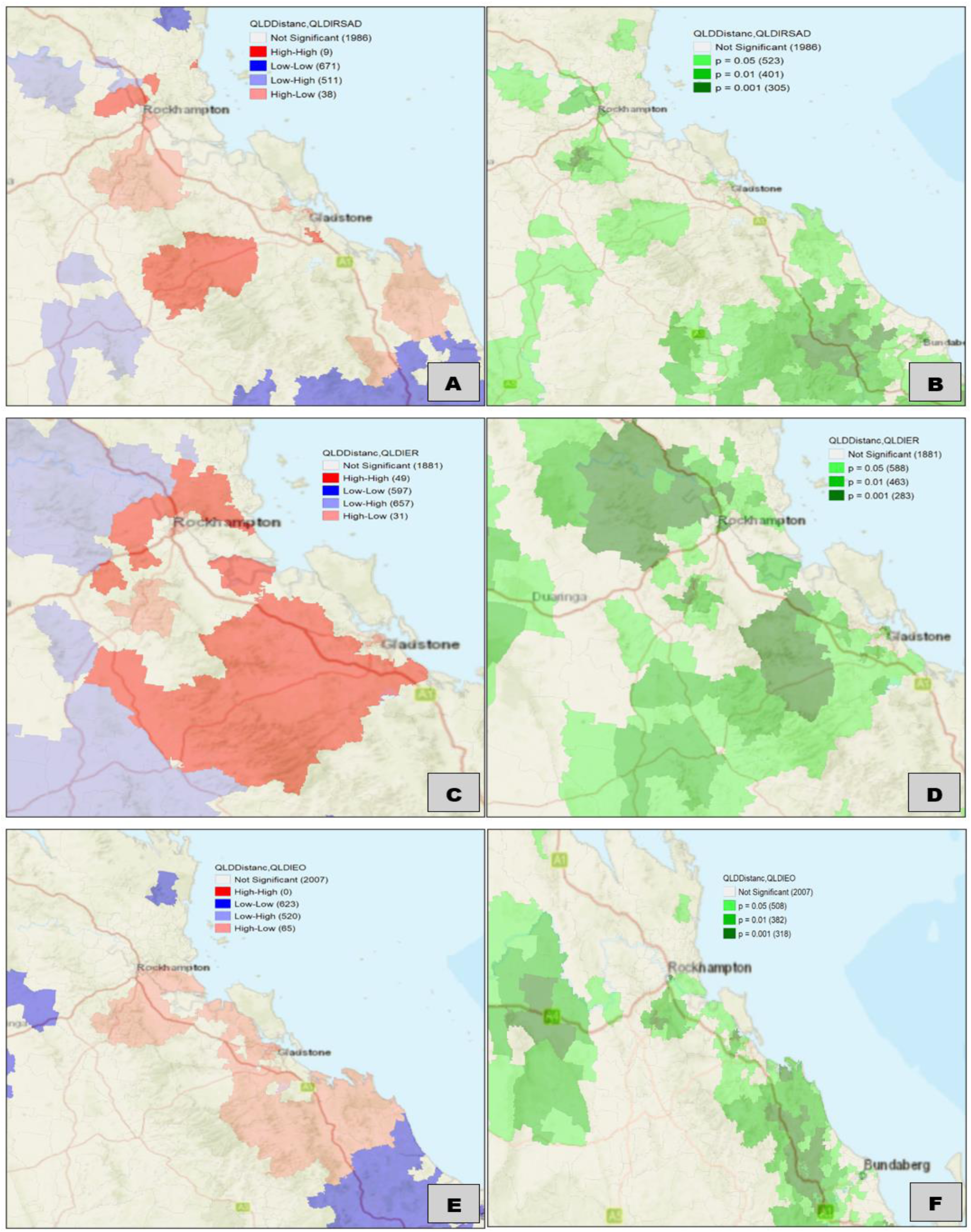
Bivariate Local Moran’s I analysis – Cluster map (left) and significant map (right) was generated from the 2016 SEIFA data, using proximity to LNG port (Distance) and the three SEIFA.

## Discussion

The aim of the study was to examine if the rate of asthma hospitalisation is associated with the proximity to an LNG terminal, through the use of spatial autocorrelation analysis and distribution analysis. Additionally, the three SEIFA were examined in this study to better understand if socioeconomic factors determined through the three SEIFA can help to better understand the role of socioeconomic position and its ability to determine the place of residence for the studied population. In the initial stage of the analysis, through data visualisation, there was evidence to suggest that the rate of asthma hospitalisation is correlated with the proximity to an LNG terminal. Suggesting that suburbs located in closer proximity to an LNG terminal, will likely record higher rates of asthma hospitalisation. Furthermore, the prevalence of asthma hospitalisation is also higher in the female group, which suggest that females is at higher risks of asthma hospitalisation than that of the male group. Figure 7 further support this finding as the distribution in rate of asthma hospitalisation is much higher amongst the female population residing in suburbs that is in closer proximity to the LNG ports. For rate of asthma hospitalisation amongst the suburbs located within the 100km buffer, one suburb consistently recorded higher rates of asthma hospitalisation in four out of the five years that this study was conducted on. Gladstone Central, a suburb located roughly 10km from the LNG terminal with a population of roughly 1546 people, consistently recorded higher rates of asthma hospitalisation compared to other neighbouring suburbs. The higher rates of asthma hospitalisation seen in Gladstone Central remains unchanged amongst the female and male groups. Gladstone Central is also the suburb with the highest number of populations, when compared to neighbouring suburbs whom of which has less than 100 people.

To further explore the spatial autocorrelation of the dataset, two spatial autocorrelation analysis were carried, and this was the Global Bivariate Moran’s I and the Bivariate Local Moran’s I. The Global Bivariate Moran’s I was utilised to identify is there is significant evidence of spatial autocorrelation amongst the whole dataset whilst the Bivariate Local Moran’s I was utilised to assess spatial autocorrelation amongst neighbouring suburbs for values that is both similar and/or dissimilar. The results obtained from these two analyses suggest that the further away a suburb is to an LNG terminal, the prevalence of asthma hospitalisation increases. Additionally, the socioeconomic position of the population within the 100km buffer was studied. The Index of Relative Socio-Economic Advantages and Disadvantages (IRSAD) shows that those residing closer to the LNG terminal is commonly those in decile 5 and decile 2. According to the ABS SEIFA decile system, the lower the decile the much more disadvantaged they were. The SEIFA decile is analysed for suburbs with records of asthma hospitalisation and not for the whole population.

Those in deciles 5 and deciles 2 on the IRSAD tends to live in closer proximity to the LNG terminals, which suggest that areas around 7km to 22km of the LNG terminals are populated with residences from a mixed socio-economic advantaged and disadvantaged background. A potential explanation can be found in population numbers, as suburbs within 7km and 22km of the LNG terminal only recorded a small number of populations, with Gladstone Central holding the greatest population count. A Bivariate Local Moran’s I analysis was carried out for the three SIEFA and their relativity to the proximity of an LNG terminal. The results show that clustering patterns varies across the three SEIFA, with less clustering seen in the IRSAD analysis (figure 10a), which is supported by the significant map (figure 10b). Therefore, according to the Bivariate Local Moran’s I analysis for the three SEIFA, the evidence suggests that the populations residing in closer proximity to the LNG terminals is less socioeconomically disadvantaged and those residing further away from the LNG terminals is much more socioeconomically disadvantaged. A possible reason to such findings may be due to housing affordability and employment opportunities in coastal areas.

In summary, the overall findings in this study suggest that suburbs that is in closer proximity to an LNG terminal, is unlikely to record higher rates of asthma hospitalisation. Additionally, those residing closer to the LNG terminals is less socioeconomically disadvantaged. A possible reason for such finding may be due to several factor. The first factor may potentially be due to type of data utilised in this analysis, specifically the asthma hospitalisation data. The use of asthma hospitalisation data generated from electronic health records may have indirectly produced a bias outcome, as there is an increase potential for missing data and misclassification of data that is crucial in providing demographic information and disease status (Conderino et al., 2024; Delfino et al., 1993; Kruse et al., 2018). The analysis method employed in this study can also be another factor that may impact the findings in this study. The non-spatial correlation outcome using bivariate spatial autocorrelation analysis suggests that alternative analysis methods should be employed to address sampling bias because of utilising bivariate analysis rather than multivariable analysis (Kim et al., 2018; Shirota & Gelfand, 2022).

In combination to the use of a multivariable analysis to address the sampling bias that may arise due to the use of a bivariate spatial autocorrelation, is the testing of confounding factors. The impact of confounding factors like air pollution may cause or exacerbate asthma symptoms leading to increase hospitalisation risks (Bronte-Moreno et al., 2023; Tiotiu et al., 2020). Air pollution data such as fine particulate matter (PM)(Li et al., 2003), industrial emissions (Sims et al., 2020) and flaring practices at natural gas production areas (Willis et al., 2020), is possible variables to incorporate into a multivariable analysis as they increase the robustness of the model and increase the statistical quality of the study. To further model the true disease burden of asthma relative to the proximity of an LNG terminal, the use of climate data like temperature (Qiu et al., 2015), rainfall (Wang et al., 2024) and ozone exposure (Zu et al., 2018) is recommended as a future study direction for spatial and spatiotemporal autocorrelation analysis (Cortes-Ramirez et al., 2024).

## Limitations

This study have potential limitations. The limitation of sample size may affect the estimates produced by the spatial autocorrelation model utilised in this study. Asthma hospitalisation rates is generated from asthma hospitalisation case numbers reported by Queensland Health and therefore do not include asthma prevalence for non-hospitalised patients, and this may limit the potential correlations of asthma prevalence and proximity to LNG terminals. Additionally, due to lack of previous research conducted on this study topic, sampling methods may have been constrained to data availability. Confounding factors like impact of fine particulate matter (PM), temperature, rainfall, and other demographic factors was not considered or factor into the study design. This was due to time constraints and data availability for PM, temperature and some demographic data. Therefore, the finding in this study may be conservative and potentially underestimate the full impact to asthma patients relative to their proximity to LNG terminals, as (1) data is only collected based on asthma hospitalisation; (2) evaluation for rate of asthma hospitalisation was examined based on population numbers, and do not take into account the suburb size or how population density per suburb is distributed relative to the LNG terminal; (3) census information regarding population size and SEIFA was collected based on the 2016 census, and therefore do not account of annual differences. A potential future direction for this study is to incorporate several confounding factors and variables in the spatial autocorrelation model to better model the full potential of the findings as multivariable analysis for spatial autocorrelation will generate a more robust dataset that will increase the quantitative significance of the finding. Despite these limitations, the findings may be interpreted with the limitation in mind as an incentive to further the study topic in order to not underestimate the true significance of asthma hospitalisation and the proximity to the LNG terminals.

## Conclusions

In conclusion, the findings in this study have identified that the lack of diverse variable incorporation or multivariable analysis can greatly impact the implementation of spatial and temporal analyses in epidemiology. Despite limitations in accessing high spatial and temporal resolution data for epidemiological analyses, the use of spatial statistics is recommended to be incorporated into future studies as spatial analysis can provide a more comprehensive visualisation of high-risk areas. Multivariable spatial analysis has great potential for prevalence modelling in complex scenarios when multiple environmental factors can be potential determinants of diseases in the general population.

## Data Availability

All data produced in the present study are available upon reasonable request to the authors

